# ASSISTED REPRODUCTIVE TECHNOLOGIES (ART) EQUITY, JUSTICE AND AUTONOMY IN GHANA

**DOI:** 10.1101/2024.10.19.24315805

**Authors:** Francis Jojo Moses Kodzo Damalie, Charles Mawunyo Senaya, Elikplim Adzo Damalie, Herbert Ekoe Dankluvi, Millicent Osaah, Beatrice Yeboah, John Jude Annan, Ellis Fleischer Djoleto, Rudolf Kantum Adageba, Alexander Tawiah Odoi

**Author notes:** Corresponding author (FJMKD).

## Abstract

Restrictive legislation, which is the main barrier to some assisted reproductive technology (ART) services in many countries, is non-existent in Ghana. However, ART services are concentrated in the capital cities of only four out of the sixteen regions, serving predominantly middle- and upper-class individuals. There is limited evidence about the factors preventing broader access to ART services in Ghana, and this study aims to document these barriers. A cross-sectional survey was conducted in July 2024 across all 22 fertility centers in Ghana, using two structured questionnaires administered via Google App to 61 ART personnel and 104 treatment defaulters. Results showed that mentorship from senior colleagues (65.57%) was the most common way for ART professionals to acquire skills. Almost all (91.80%) professionals offered a full range of ART procedures, but 86.89% advocated for regulated practice. They identified high treatment costs (70.49%) and lack of awareness (16.39%) as the most significant barriers. Among treatment defaulters, 88.47% had sought ART services based on word-of-mouth recommendations, compared to only 4.8% influenced by traditional or social media. More than half (50.96%) of the women were in their thirties, and 48.08% required in vitro fertilization (IVF). While 58.65% sought treatment within five years of infertility, 70.2% discontinued due to high costs, and 35.57% due to partner non-availability. Despite the absence of restrictive policies for ART services in Ghana, Prohibitive costs, partner non-availability, and lack of awareness limit access. However, ART professionals expressed the need for regulated practices.

## INTRODUCTION

Ghanaian families have a pro-natalist attitude and expect children to maintain family lineage, happiness, and harmony. Childbirth, therefore, guarantees marital stability and economic or social security in a society that lacks formal social security systems for older people. Infertility, thus, exerts devastating biomedical, psychological and sociocultural consequences on affected couples [1, 2]. The exact burden of infertility in Ghana is not known, but it is estimated to affect about 11.8% of women and 15.8% of men [3].

Tubal and severe male factors are the commonest causes of Infertility in Ghana, for which assisted reproductive technology (ART) treatment is the most effective. Many Ghanaians are turning to ART treatment for assistance to achieve their dream of parenthood. ART refers to medical procedures that involve the manipulation of eggs, sperm or embryos outside the body to facilitate conception. The typical approaches are in-vitro fertilisation and intra-cytoplasmic sperm injection. Ghana requires about 1500 ART cycles per 1 million people a year (i.e., 45,000 IVF cycles each year for her 30 million population) for adequate fertility treatment [4, 5]. Even though statistics on IVF cycles are lacking, it is unlikely that the few ART centres in Ghana can provide the required number of IVF cycles. For instance, in 2020, four out of the fifteen ART centres in Ghana reported 677 cycles [6]. This suggests a high unmet need for IVF services.

While restrictive legislation and regulation appear to be the main barriers to infertility treatment in developed countries [7], the factors limiting access to ART services in Ghana are poorly documented. Some studies found that the high cost of setting up and associated scarcity of ART centres, the high cost of ART treatments, socioeconomic deprivation, and the lack of skilled infertility personnel make ART largely inaccessible in Africa [8-11]. Understanding the landscape of ART access in Ghana is crucial for policymakers, healthcare providers, and researchers to address existing barriers and improve the availability and affordability of fertility treatments for all individuals and couples in need. Thus, this study seeks to document the barriers to ART services in Ghana.

## Materials and Methods

This cross-sectional study was conducted across twenty-two fertility centres in Ghana from 1^st^ to 31^st^ July 2024. A structured questionnaire was administered via a Google app to 61 ART personnel from all fertility centres in Ghana through the Fertility Society of Ghana (FERSOG) WhatsApp group to obtain information on fertility professionals’ sociodemographic characteristics, training, work experience and the type of services they offered. Another structured questionnaire was administered to patients who defaulted treatment at Hallmark Medicals, a private fertility centre in Kumasi, through telephone conversations. The patients were conveniently identified through the hospital’s records and contacted to obtain information on their knowledge of ART services and reasons for dropping out of the recommended treatment. Ethical approval CHRPE/AP/205/24 was obtained from the Committee for Human Research Publication and Ethics, Kwame Nkrumah University of Science and Technology (KNUST). Written informed consent was obtained from each participant before enrolling in the study. Data were analysed using STATA version 14.0.

## Results

### Demographics Characteristics of ART Healthcare Professionals in Ghana

Table 1 shows the demographic characteristics of ART professionals in Ghana. Most of the personnel, 39(63.93%) were males. Thus, the gender distribution of the personnel was almost two males to one female. The majority of the personnel, 43(70.49%), were aged between 30 and 49. Embryologists constituted nearly a third of the health professionals’ group, with 18 members making up 29.51% of all healthcare personnel, followed by fertility nurses 17(27.87%). There was only one clinical psychologist among the ART personnel.

**Table 1:** Demographic Characteristics of ART Healthcare Professionals in Ghana.

| Variables | Frequency<br>(n=61) | Percentage |
| --- | --- | --- |
| Sex |  |  |
| Female | 22 | 36.07 |
| Male | 39 | 63.93 |
| Age (Years) |  |  |
| < 30 | 8 | 13.12 |
| 30-39 | 24 | 39.34 |
| 40-49 | 19 | 31.15 |
| 50-59 | 9 | 14.75 |
| >60 | 1 | 1.64 |
| Clinical Role |  |  |
| Reproductive Endocrinology Sub-specialist | 8 | 13.11 |
| Obstetrician and Gynaecologist | 14 | 22.95 |
| Embryologist | 18 | 29.51 |
| Fertility Nurse | 17 | 27.87 |
| Clinical psychologist | 1 | 1.64 |
| Other (Counselor, Urologist, and Fellow in training) | 3 | 4.92 |
| **Level of Education** |  |  |
| Clinical Fellowship | 12 | 19.67 |
| PhD | 3 | 4.92 |
| Membership | 11 | 18.03 |
| Postgraduate Masters | 9 | 14.75 |
| Postgraduate Diploma | 1 | 1.64 |
| MBChB | 2 | 3.28 |
| Degree in Biomedical Sciences, Laboratory Technology | 4 | 6.56 |
| Degree in Nursing/Midwifery | 9 | 14.75 |
| Degree in Psychology | 1 | 1.64 |
| Diploma in Nursing/Midwifery | 4 | 6.56 |
| Diploma in Perioperative Nursing | 1 | 1.64 |
| Nurse Assistant- Clinical (NAC) | 3 | 4.92 |
| Other (Degree Chemistry) | 1 | 1.64 |

Clinical Fellowship and membership were the most common and highest educational levels attained by ART professionals, followed equally by postgraduate masters and degrees in midwifery. Five per cent of the personnel had a PhD.

### Training and Experience in ART

Table 2 shows the survey results on training ART professionals and their constraints. Training on the Job (Mentorship) emerged as the most frequent approach used by 20(32.79%) ART professionals to acquire knowledge and skills, followed by Clinical Fellowship Training 12(19.67%). Notably, 3(4.92%) of ART professionals had a PhD. Interestingly, two out of three 40(65.57%) respondents received ART training in Ghana. Aside from Ghana, India and Europe were featured as favourite destinations for ART training. Most professionals, 41(67.22%), reported having less than five years of experience in ART.

**Table 2:** Training and Experience in ART.

| **Variables** | **Frequency**<br>**(n=61)** | **Percentage** |
| --- | --- | --- |
| **Level of Professional Training in ART** |  |  |
| Fellowship | 12 | 19.67 |
| PhD | 3 | 4.92 |
| Post Graduate Masters | 7 | 11.48 |
| Post Graduate Diploma | 5 | 8.20 |
| Six Months Observership | 3 | 4.91 |
| Training on the Job (Mentorship) | 20 | 32.79 |
| Other (Senior Resident, Fellow in Training) | 11 | 18.03 |
| **Country of Training in ART** |  |  |
| Europe | 5 | 8.20 |
| South Africa | 2 | 3.28 |
| Australia | 1 | 1.64 |
| India | 12 | 19.67 |
| Ghana | 40 | 65.57 |
| Other (Nigeria) | 1 | 1.64 |
| **Challenges in ART training** |  |  |
| Difficulty financing training; Difficulty finding mentors | 19 | 31.15 |
| Difficulty with traveling abroad for training; Difficulty financing training; Few opportunities available for training; Difficult finding a mentor | 41 | 67.21 |
| Difficulty with traveling abroad for training; Difficulty finding mentors | 1 | 1.64 |
| **Duration of ART Experience (years)** |  |  |
| <1 | 6 | 9.84 |
| 1-5 | 35 | 57.38 |
| 6-10 | 14 | 22.95 |
| >10 | 6 | 9.83 |
| **Restrictive and Regulated ART Environment** |  |  |
| Yes | 53 | 86.89 |
| No | 8 | 13.11 |

### ART services offered and their barriers

An overwhelming majority of ART professionals had been involved in or offered almost all ART services available globally, including IVF for single women, commercial surrogacy, multifetal pregnancy reduction, intrafamilial donor sperm insemination and sex determination. Only 2(3.28%) of ART personnel had never been involved or offered IVF for single women, multifetal pregnancy reduction and sex determination (Table 3).

**Table 3:** Types of ART services offered by ART professionals.

| Variable | Frequency | Percentage |
| --- | --- | --- |
| **Services involved in or offered.** |  |  |
| IVF for single women; Use of donor gametes; Commercial surrogacy; Embryo adoption; Multifetal pregnancy reduction; Cryopreservation and transfer of frozen-thawed embryo (FET); intra-familial donor insemination; Sex selection; pre-implantation genetic diagnosis (PGD) | 56 | 91.80 |
| Use of donor gametes; Commercial surrogacy; Embryo adoption; Pre-implantation genetic diagnosis (PGD); Cryopreservation and transfer of frozen-thawed embryo, (FET) and Intra familial donor insemination | 2 | 3.28 |
| Sex selection | 3 | 4.92 |

Over 70% of ART personnel mentioned the high cost of services as the main barrier to ART in Ghana, followed by poor knowledge of the availability of ART services by the patients 10 (16.39%). Only an insignificant 6.56% of ART professionals considered religion as a barrier to ART (Fig1).

**Fig 1:**
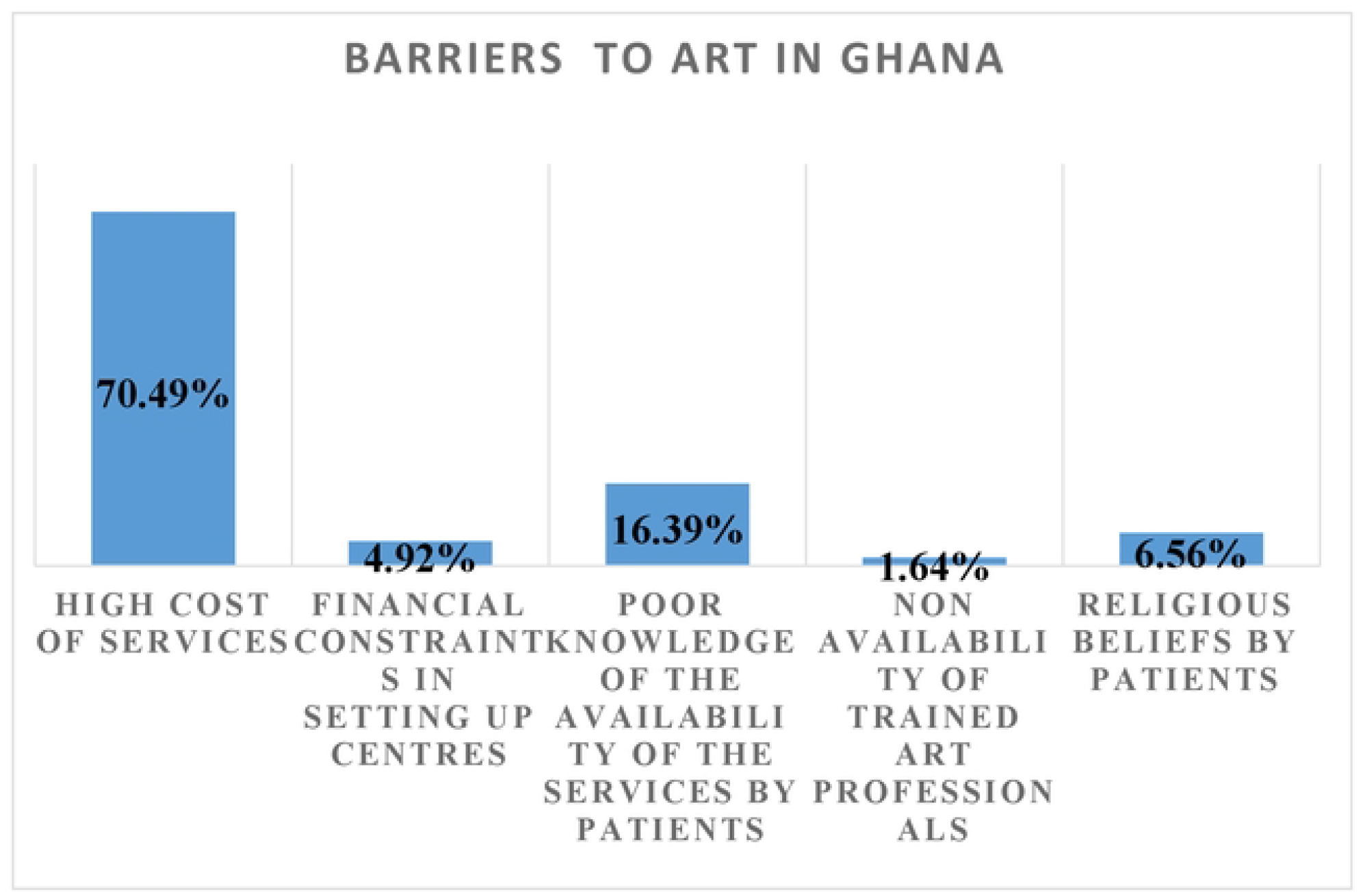
Barriers to ART services identified by ART professionals

### Demographic Characteristics of Women Who Defaulted in Infertility Treatment

Fig 2, 3 and 4 show the demographic characteristics of 104 fertility clinic attendants who defaulted in treatment. More than half 53(50.90%) of these women were in their thirties (30-39 years), and about a third, 32(30.77%), were in their forties (Fig 2). The overwhelming majority of the defaulters, 90(86.54%) were Christians (Fig 3) and 91(87.50%), were married (Fig 4). Only a very small proportion, 5(4.81%) of these women who desired a pregnancy did not have a partner (Fig 4).

**Fig 2:**
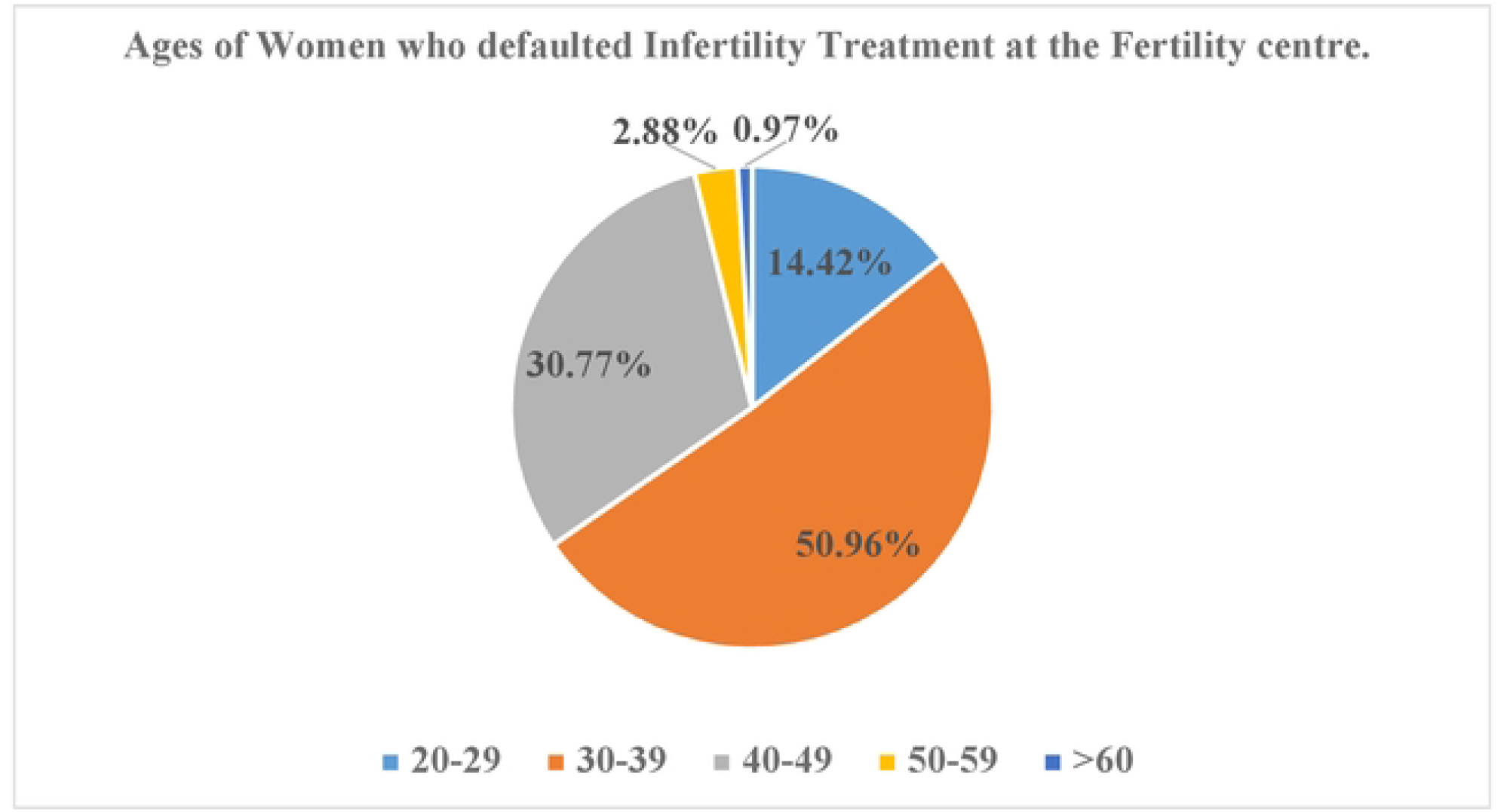
Age distributions of ART clinic attendants who defaulted treatment

**Fig 3:**
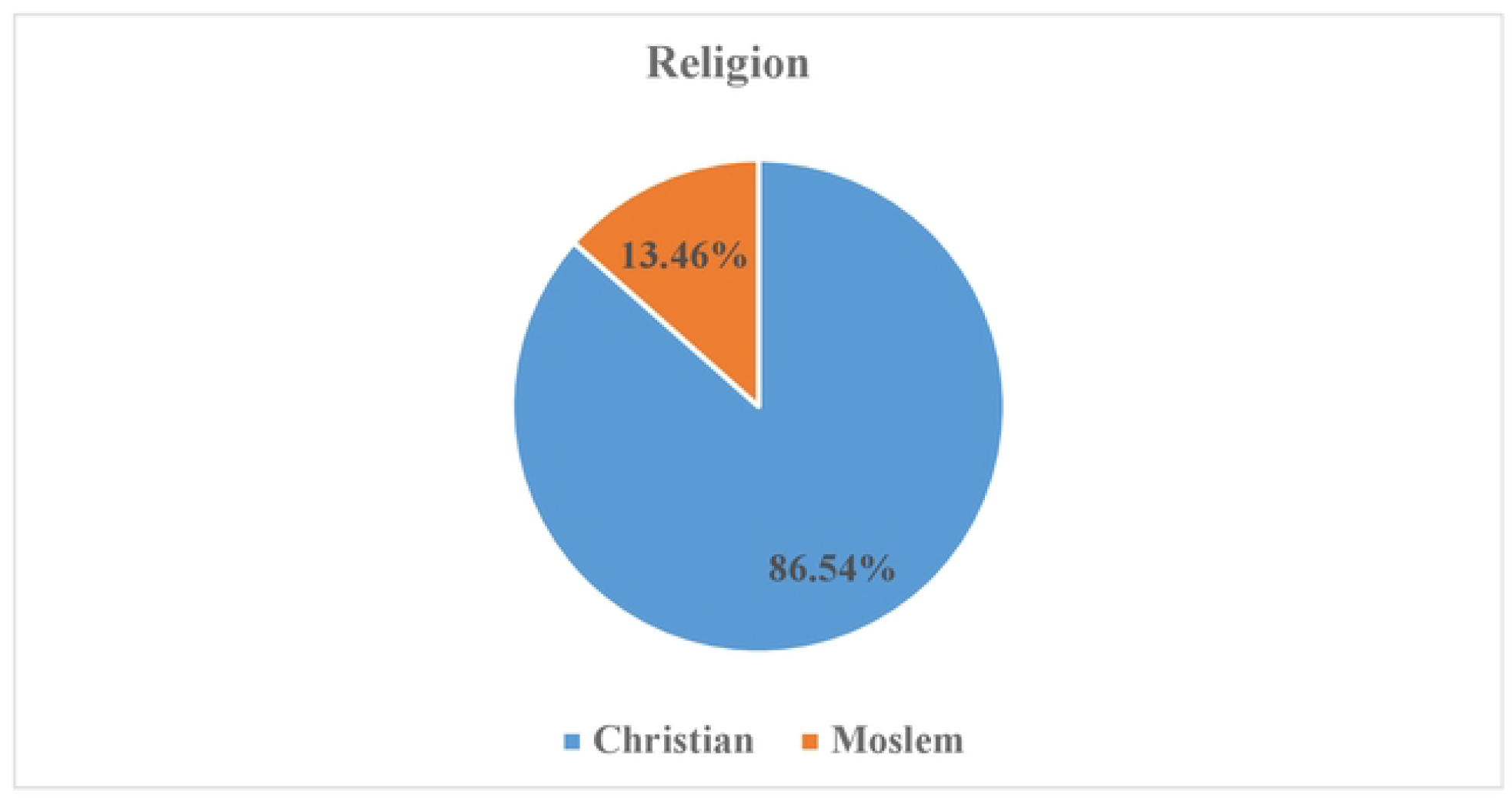
The distribution of ART clinic defaulters according to religion

**Fig 4:**
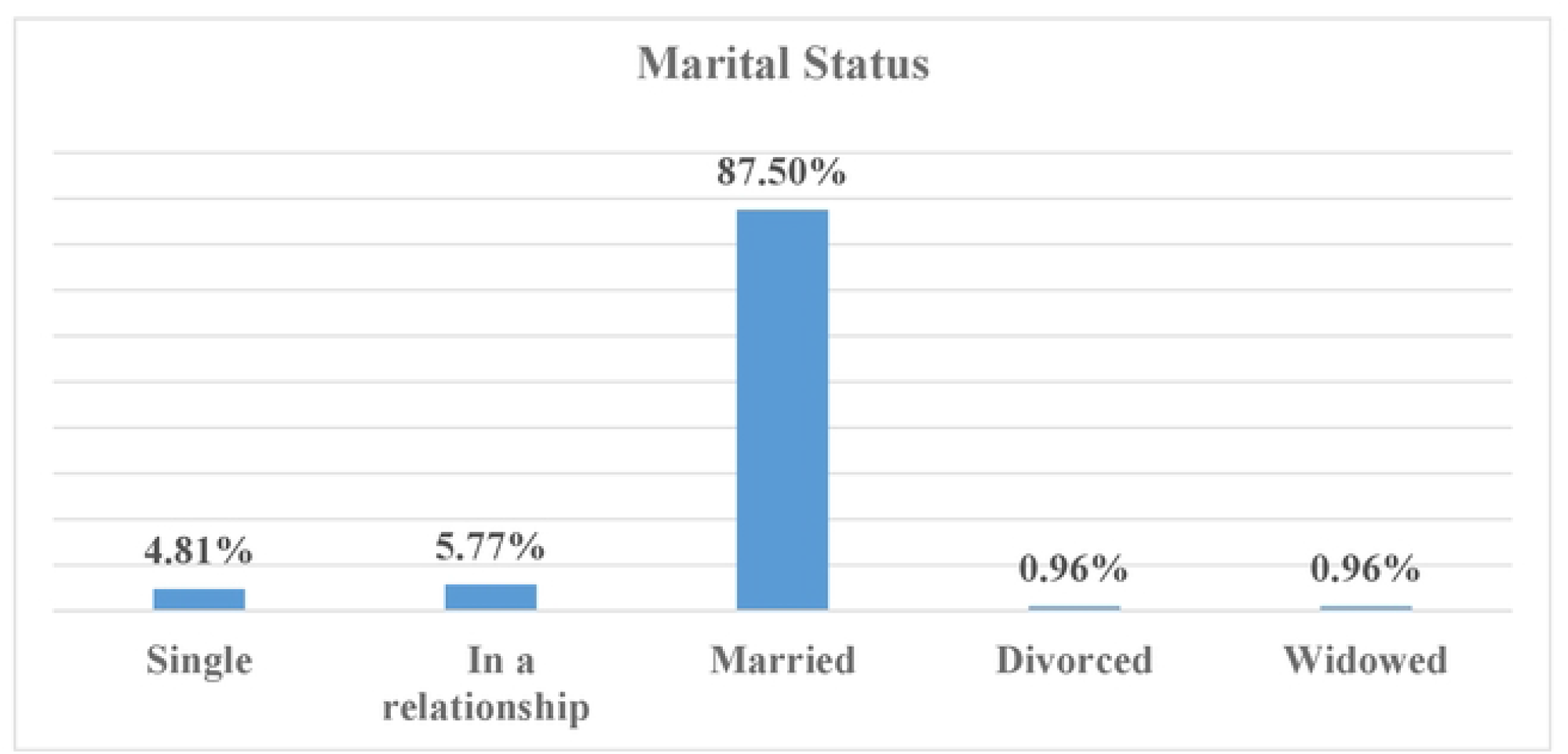
The distribution of ART clinic defaulters according to marital status

### Living experiences of women with infertility and their sources of referral for ART

Table 4 shows the living experiences of the women with infertility and their sources of referral for ART. Forty-five per cent (45.19%) of these defaulters sought treatment after 2-5 years of Infertility. About a quarter of them did so after 6-10 years. Almost two-thirds, 74 (71.15%) of the couples, live together. Nearly a third of the couples were in distant relationships, and among these, 13(43.34%) had their partners abroad, while more than a third lived in different cities in Ghana.

**Table 4:** Living experiences of the women with infertility.

| Variables | Frequency<br>(n=104) | Percentage (%) |
| --- | --- | --- |
| **Duration of Infertility (Years)** |  |  |
| <1 | 14 | 13.46 |
| 2-5 | 47 | 45.19 |
| 6-10 | 26 | 25.00 |
| 11-20 | 12 | 11.54 |
| >20 | 5 | 4.81 |
| **Couples living together?** |  |  |
| Yes | 74 | 71.15 |
| No | 30 | 28.85 |
| **Residence of partner if couples don't live together** |  |  |
| Abroad | 13 | 43.34 |
| In Ghana, but in different cities or town | 11 | 36.66 |
| In the same city but in different locations | 6 | 20.00 |
| **Source of information about the ART clinic** |  |  |
| Recommendation from a friend, family or colleague who has patronised the services | 36 | 34.62 |
| Recommendation from a friend or family who has heard about the hospital but has not patronised it themselves | 56 | 53.85 |
| Radio | 2 | 1.92 |
| Hospital social media handle | 3 | 2.88 |
| Others (Nurse, Doctor, and Sign board) | 7 | 6.73 |

Word-of-mouth recommendations from people who had a personal experience with ART or knew someone with the experience was the source of referral to the ART clinic in an overwhelming 88.47% of instances. Radio and social media, combined, constituted an insignificant 4.88% of sources of information on the ART clinic. Health professionals were the sources of information in another 6.73% of instances.

### Causes of Infertility and Fertility Treatments Offered and Received

Fibroids was the commonest diagnosis made in one out of four 27(25.95%) of these women with Infertility who defaulted treatment, followed by unexplained Infertility in one out of five (20.19%) and tubal blockage 15(14.44%). Male Infertility and anovulation contributed almost equally, 9(8.65%) vs 10(9.62%) to the Infertility in these women. In nearly one out of five cases (17.30%), the cause of Infertility was any combination of advanced age, anovulation, tubal factor, male factor and fibroids.

In-vitro fertilisation (IVF) was the most common treatment offered to almost half (48.08%) of these women with Infertility, followed by ovulation induction with timed intercourse in about a third, 35(33.65%), and ovulation induction with intra-uterine insemination (IUI) in 14(13.46%) of the cases. Intra-cytoplasmic sperm injection was only infrequently offered in 3(2.88%) of cases. Very few of these women, only 14.42%, had had any form of infertility treatment, ovulation induction and timed intercourse being the commonest treatment received by about half of these women, while just more than a quarter each had received either ovulation induction with IUI or IVF treatment (Table 5).

**Table 5:** Causes of infertility and treatments offered and received.

| Variables | Frequency<br>(n=104) | Percentage (%) |
| --- | --- | --- |
| **Cause of the Infertility** |  |  |
| Abnormal semen parameters | 9 | 8.65 |
| Advanced age | 4 | 3.85 |
| Anovulation | 10 | 9.62 |
| Fibroids | 27 | 25.95 |
| Tubal blockage | 15 | 14.44 |
| Unexplained | 21 | 20.19 |
| Advanced age; Anovulation; Tubal blockage;<br>Fibroid | 9 | 8.65 |
| Tubal blockage; Abnormal semen parameters;<br>Unexplained | 9 | 8.65 |
| **Treatment Offered** |  |  |
| In-vitro fertilisation (IVF) | 50 | 48.08 |
| Intra-Cytoplasmic Sperm Injection (ICSI) | 3 | 2.88 |
| Ovulation induction and intrauterine<br>insemination | 14 | 13.46 |
| Ovulation induction and timed intercourse | 35 | 33.65 |
| Ovulation induction and timed intercourse; In-Vitro Fertilization (IVF) | 2 | 1.93 |
| **ART Treatments received before** |  |  |
| Yes | 15 | 14.42 |
| No | 89 | 85.58 |
| **ART Treatments Received if "Yes"** |  |  |
| Ovulation induction and timed intercourse | 7 | 46.66 |
| Ovulation induction and intrauterine<br>insemination (IUI) | 4 | 26.67 |
| In-Vitro Fertilization (IVF) | 4 | 26.67 |

### Follow Up and Reasons for Defaulting Treatment

Two out of five defaulting patients mentioned the high cost of treatment, while one out of five cited partner non-availability as the reason for default. The high cost of treatment and partner non-availability were the reasons for default among 14.42% of the patients. Thus, the high cost of treatment stood out as the commonest reason for defaulting to ART treatment (Table 6).

**Table 6:** Follow-up and reasons for not returning.

| Variables | Frequency<br>(n=104) | Percentage<br>(%) |
| --- | --- | --- |
| **Last visit to the Clinic (Months)** |  |  |
| <6 | 39 | 37.50 |
| 7-12 | 42 | 40.38 |
| 13-24 | 13 | 12.50 |
| >24 | 10 | 9.62 |
| **Reason for not returning for the treatment recommended** |  |  |
| My partner is not available for the treatment | 19 | 18.27 |
| We can afford the treatment, but we do not like the care at the clinic | 1 | 0.96 |
| The clinic is too far from where we live | 5 | 4.81 |
| The cost of treatment is too high | 44 | 42.31 |
| We already have a child or children and are no longer keen on the treatment | 3 | 2.88 |
| The cost of treatment is too high; My partner is not available for the treatment | 15 | 14.42 |
| The cost of treatment is too high; We already have a child or children and are no longer keen on the treatment | 5 | 4.82 |
| The cost of treatment is too high. We have found another place where we prefer to have the treatment | 9 | 8.65 |
| My partner is not available for the treatment.; The clinic is too far from where we live | 3 | 2.88 |

## Discussion

We interviewed 61 health professionals; more than half (52.46%) were less than 40 years old, just a little more than a third (36.06%) were clinicians (Obstetricians and Gynaecologists/Reproductive Endocrinologists and Infertility experts), about a third (29.51%) were embryologists and the remaining third (34.43%) were fertility nurses and other supporting staff such as counsellors or clinical psychologists. In a similar study by Whittaker et al. on access to ART in sub-Saharan Africa, a smaller sample size of 31 participants was involved [12]. While their research primarily included only fertility specialists and embryologists, ours covered a broader participant base to include the complete complement of healthcare professionals involved in ART. About two-thirds of the health professionals acquired their ART subspecialty skills through Fellowship and various postgraduate studies (PhD, postgraduate masters and diplomas), while a significant third (37.7%) did so through mentorship or observership with more experienced senior colleagues. Very importantly, a majority (65.57%) of professionals acquired their skills locally in Ghana. Among the 34.43% who had their training abroad, India was the most common destination visited by 19.67%, followed by Europe (8.20%). These findings were similar to observations in Southeastern Nigeria, where all ART practitioners, primarily obstetricians/gynaecologists, received their training from India [13] [14]. Most ART professionals interviewed were inexperienced because a majority (57.38%) of respondents had less than five years in ART practice. Even though this may appear less assuring of the quality of ART services in Ghana, the young age of most professionals offers the opportunity for a more extended and vibrant ART service.

Advocacy by the majority (86.89%0 of ART professionals for a restrictive environment for the practice speaks of members of a fraternity who are intent on curbing the excesses of the practice. The ease of access to all services, including IVF for single women, commercial surrogacy, multifetal pregnancy reduction, intrafamilial donor insemination, and sex selection, suggests that couples with Infertility in Ghana have high patient autonomy. Only 3.28% of professionals had not offered or been involved in IVF for single women, multifetal pregnancy reduction and sex selection. Future studies must establish why some ART experts have never provided or been engaged in offering these services. [15] [16].

For more than two-thirds (67.21) of ART professionals, a combination of difficulties associated with travelling abroad, financing the ART training, finding training opportunities, and finding a mentor were the main challenges to training. These findings were consistent with reports on ART in Africa, which identified the lack of structured training and financial constraints as factors militating against ART training [12]. The African network and registry for ART report for 2020 further underscores the need for accessible and affordable training to address these gaps [15]. Mentorship was significant for transferring ART knowledge and skills, contributing to training a third (32.79%) of all professionals in this study. [17]. At the time of collecting this data, the Reproductive Endocrinology and Infertility (REI) unit of the Ghana College of Physicians and Surgeons had just admitted its first-year Fellows. The Ghana College of Nurses and Midwives also took their first year of Membership students in Fertility Nursing this year (2024). The Ghana Association of Embryologists (GACE) was in talks with stakeholders about starting formal professional training for embryologists. This formal training of ART professionals is expected to improve the number of professionals with ART subspecialty skills. The opportunities these colleges offer will obviate the need to travel abroad for studies and the attendant financing burdens.

Almost three-quarters (70.49%) of ART professionals identified high ART treatment costs, while 16.39% identified the lack of awareness for ART services as the central berries for treatment. These findings are in consonance with Whittaker et al.’s study on access to ART in sub-Saharan Africa, where high treatment costs, lack of public funding, poor policy awareness, and a shortage of skilled professionals were major obstacles [12]. The high cost and uneven distribution of ART services in Ghana’s burgeoning cities create inequities concerning who can afford or access the services. Appropriate government interventions, such as tax exemptions and regulations on the cost of ART drugs and consumables, can significantly mitigate the high cost associated with both access for clients and the setup of new clinics.

A majority of the women who defaulted treatment at the centre were married (87.50%), Christian (86.54%), young women, 65.38% being below 40 years of age. These findings align with other observations in which most women utilising ART services were in their thirties [18-20], identified as Christians [19, 21] and were mostly married [19-21]. We postulate that the default rate was higher among the younger women either because of an exaggerated confidence in fertility or because they were more dependent on their partners for the final decision and finances for the treatment.

Over half (58.65%) of the treatment defaulters sought treatment early, within five years. These observations were similar to those of a study in which the majority (40%) of women utilising ART services in some selected facilities in Accra had experienced Infertility in less than five years, followed by 35.33% in 5-9 years [22]. In another study which explored the experiences of women accessing ART services in Ghana, it was found that most women had been married for years, had tried natural conception and eventually resorted to ART services because they were either worried about their age or desired to satisfy their spouses and avoid the displeasure of in-laws [20].

Word-of-mouth recommendation was a more important referral source to the clinic than traditional and social media. This is because an overwhelming 88.47% of these treatment defaulters visited the clinic based on word-of-mouth recommendations compared to only 4.8% who did so based on traditional and social media adverts. This means that satisfied clients were more prone to return with another patient. Word-of-mouth was also the primary source of information about ART for women with infertility seeking ART services in Northern Nigeria [18], with family relations giving the most information (46.0%), followed by friends (28.7%) and health facilities (18.0%).

Distant relationships featured as a possible contributor to infertility among these treatment defaulters because a significant three out of ten couples do not live together. Among these, 43.34% had their partners abroad.

One out of four women in this study had fibroids identified as the cause of Infertility. Fibroids are prevalent among women with infertility [23, 24], and their prevalence as a cause of Infertility among ART treatment defaulters in this study corroborates existing literature on the high prevalence of fibroids among African women [25]. Fibroids cause Infertility through impaired uterine contractility, altered endometrial receptiveness [26], blockage of fallopian tubes and prevention of gamete passage [27]. Unexplained Infertility was the second most common cause of Infertility identified, responsible for infertility among 20.19% of these women. This compares with the findings in a study in Zimbabwe, which reports unexplained infertility as the most common cause of Infertility, affecting 22% of the women seeking treatment at gynaecological clinics [28]. Unexplained infertility is a diagnosis of exclusion for couples who are unable to conceive despite regular unprotected sex and do not fit the criteria for diagnosis of male or female factor infertility [29]. In various reviews regarding infertility, unexplained infertility has been reported to account for varying numbers of cases, ranging between 10.4% - 30% [30-32]. Tubal blockage may be caused by sexually transmitted infections, pelvic infections, fibroids, pelvic adhesions after fibroid surgery and other factors. It is reported to account for 30-40% of female infertility. Where resources and skills are available, it may be managed by tubal microsurgery. However, the majority of case management requires IVF [33]. In the present study, it was the third most common cause of infertility, accounting for 14.44% of infertility among women defaulting to fertility treatments at the centre. This finding is consistent with data from other regions where tubal factors are a common cause of infertility. For instance, in Zimbabwe, tubal blockage was responsible for 20% of infertility among women seeking fertility treatment [28]. Similarly, it represented 22.3% and 25.5% of the causes of infertility cases among Omani women [34] and women in Austria [35], respectively. It was also observed that male infertility contributed to 8.65% of infertility cases in this study. This proportion of male factor contribution is low compared with that in Zimbabwe (19%) and reports across Africa (22.26%). The observed discrepancy could be because many patients presented semen analysis from non-standardised laboratories, where the WHO strict criteria were not applied.

In-vitro fertilisation (IVF) was the most common treatment offered (48.08%) to these women who defaulted, followed by ovulation induction with timed intercourse (33.65%). In-vitro fertilisation, ovulation induction with timed intercourse or intrauterine insemination (IUI) are common treatment options offered to couples requiring fertility treatment, even though the latter two are not assisted reproductive technologies [31]. IVF offers the best success results [36, 37]. Ovulation induction involves using drugs to trigger the release of mature oocytes. [31]. For fertilisation to occur after the induction, it can be coupled with timed intercourse or intra-uterine insemination in which washed partner spermatozoa are placed in the uterine cavity near the tubal ostia. Ovulation induction with timed intercourse is frequently used for women with less severe infertility issues, such as ovulatory disorders [36]. Furthermore, it is less invasive and costly compared with IVF, making it a first-line treatment option. In this study, only a small number, 14.42%, had some form of treatment before presenting to the fertility centre. Almost half (46.66%) of them had had ovulation induction with timed intercourse, suggesting that their diagnosed cause of Infertility could not have been severe.

The reasons for ART treatment defaults provided by these women in this study were the high treatment cost (70.20%) and partner non-availability (35.57%). These findings agree with those of a study in Northern Nigeria, where women receiving ART services cited high financial costs as a barrier to continued use of the services [18]. Other studies exploring the experiences and challenges of Ghanaian women using ART services reported high costs as a significant challenge for their continued fertility treatment [20, 21]. The financial burden associated with ART service utilisation is a recognised barrier to ART access in low and middle-income countries, particularly the African subregion [12, 15, 38]. The high cost of ART services in Africa has been attributed to the capital-intensive nature of setting up ART facilities, the highly privatised services, the limited number of clinics relative to demand, the high operational cost and a lack of government support [12, 15, 24]. Even though couples with infertility in Ghana enjoy autonomy in their choices of ART treatment options, this may be compromised when financial constraints limit their choices. Addressing these challenges with strategies such as introducing supportive regulatory frameworks and policies to reduce operational costs and promote public investments in ART services can make these services more accessible and affordable. Without government support or subsidised programmes, the economic burden on patients impedes equitable access.

Partner non-availability was the second most common reason for the discontinuation of ART services by 18.27% of respondents in this study. Similarly, Arhin et al. reported a lack of support from male partners, seeking alternative treatment and relocation as major reasons for discontinued ART service utilisation [39]. The non-involvement or unavailability of male partners in seeking fertility treatment is problematic because males contribute equally to Infertility and must be evaluated to arrive at a conclusive diagnosis of the cause of Infertility. Furthermore, ART treatment can be an emotionally challenging journey, making emotional support from a partner essential for the overall outcome.

## Conclusion

The findings of this study highlight significant barriers to accessing ART services in Ghana despite the absence of restrictive legislation that allows for greater patient autonomy. The primary obstacles identified were prohibitive treatment costs, lack of partner availability, and insufficient awareness of ART services. These barriers contribute to high levels of inequity and injustice in the accessibility of ART for Ghanaians facing infertility challenges. Moreover, while ART professionals in Ghana are generally well-educated and capable of providing a broad range of services, the majority desire a more regulated practice environment. To improve equity and justice for ART services in Ghana, it is essential to address these financial, informational and regulatory challenges, ensuring that all individuals in need can benefit from the available reproductive technologies.

## Data Availability

The data underlying the results presented in the study are available from the corresponding author upon request

## Acknowledgements

The authors wish to acknowledge the participants who consented and contributed to the success of this study.

